# Deep Neural Patchworks Predict Renal Imaging Biomarkers from Non-Contrast MRI via Knowledge Transfer from Arterial-Phase Contrast-Enhanced MRI

**DOI:** 10.64898/2026.02.24.26346961

**Authors:** Kai F. Kästingschäfer, Anna Fink, Stephan Rau, Marco Reisert, Elias Kellner, Janis M. Nolde, Anna Köttgen, Peggy Sekula, Fabian Bamberg, Maximilian F. Russe

## Abstract

**Rationale and Objectives:** Contrast-enhanced (CE) MRI provides clear corticomedullary contrast for renal compartment delineation but may be contraindicated or undesirable in routine practice. We aimed to enable automated extraction of renal imaging biomarkers from routine non-contrast-enhanced (NCE) T1-weighted MRI by transferring CE-derived compartment labels.

**Materials and Methods:** This retrospective single-center study (January 2017 to December 2021) included 200 participants with paired arterial-phase CE and NCE T1-weighted MRI. Cortex, medulla, and sinus were manually segmented on CE MRI and rigidly transferred to NCE MRI to provide voxel-level reference labels. A hierarchical 3D Deep Neural Patchworks model was trained on 100 examinations (90 training/10 validation) and evaluated on an independent test set of 100 examinations using the transferred CE masks on NCE as reference. Performance was assessed using Dice similarity of segmentations and biomarker agreement using volumes and surface areas (Pearson/Spearman, MAE, Lin’s CCC, and Bland-Altman).

**Results:** Whole-kidney segmentation Dice was 0.950 (left) and 0.953 (right). Total kidney volume showed high agreement with minimal bias (MAE 8.76 mL, 2.5% of mean; CCC 0.983; bias -1.56 mL; 95% limits of agreement -28.81 to 25.69 mL). Cortex volume was modestly overestimated and medulla volume underestimated, shifting predicted compartment fractions toward cortex (74.7% vs. 72,1% in ground truth; medulla 21.5% vs. 24.3%; sinus 3.8% vs. 3.6%. Sinus volume maintained high concordance despite higher Dice dispersion. Surface area was systematically underestimated with low concordance.

**Conclusion:** CE-supervised knowledge transfer enables accurate, well-calibrated kidney volumetry from routine NCE MRI and supports contrast-free renal biomarker extraction. Surface area estimation remains challenging.

**Take-home Messages:**

- CE-supervised label transfer enables accurate, well-calibrated contrast-free kidney volumetry on routine non-contrast T1-weighted MRI.
- Compartment volumetry is feasible but shows systematic cortex overestimation and medulla underestimation; surface area remains non-interchangeable due to boundary uncertainty.

## Introduction

Chronic kidney disease (CKD) is a major global health burden. In 2017, an estimated 697.5 million people worldwide were living with CKD, corresponding to a global prevalence of about 9%. [1] CKD is also among the leading causes of years of life lost worldwide and has risen substantially in global mortality rankings over the past two decades [2]. Early detection and monitoring remain challenging because standard clinical markers and imaging (for example serum creatinine/eGFR, albuminuria, and ultrasound) can be insensitive to early kidney damage. [3]

Imaging-derived structural biomarkers provide an additional window into renal integrity. In autosomal dominant polycystic kidney disease (ADPKD), total kidney volume (TKV) and its growth rate were shown to predict future functional decline in the CRISP study. [4,5] Moreover, therapies targeting cyst growth can change these trajectories: in a phase III trial, tolvaptan slowed TKV expansion and was associated with a slower decline in kidney function. [6] Beyond cystic disease, cortical volume is a key imaging component for estimating nephron number, which directly corresponds to biopsy-based glomerular density, thereby supporting its use as a marker of irreversible parenchymal loss by providing a structural correlate of nephron endowment and parenchymal integrity. [7]. Beyond direct clinical risk estimation, precise kidney volumetry in large population cohorts enables investigation of renal biology at a fundamental level, for instance through integrative analyses combining imaging with genetic and molecular data [8].

Whole-kidney segmentation relies largely on organ boundary detection, whereas compartment segmentation requires reliable internal tissue differentiation. Contrast-enhanced T1-weighted MRI provides robust corticomedullary contrast and thus a practical reference standard for delineating cortex, medulla, and sinus. However, in advanced CKD, gadolinium-based contrast agents (GBCAs) are often approached with caution; although the risk of nephrogenic systemic fibrosis is very low for group II agents, historical NSF risk and concerns about gadolinium retention continue to influence clinical practice and study design. [9,10] These considerations, along with cost and workflow constraints, limit contrast-enhanced MRI for routine follow-up and large epidemiologic studies. Non-contrast MRI is widely available, but conventional approaches can provide limited corticomedullary differentiation, making reliable manual compartment delineation difficult in many settings. [11] Automated segmentation and volumetric analysis of internal structures from native MRI has been demonstrated in small healthy cohorts using combined T1- and T2-weighted imaging. [12]

Deep learning has further improved segmentation performance across diverse MRI protocols. For kidney segmentation, U-Net style convolutional networks can achieve near human-level accuracy on non-contrast body MRI at population scale (e.g., Dice ∼0.95 for parenchymal kidney volume in UK Biobank neck-to-knee MRI). [13] Automated whole-kidney segmentation in CKD cohorts has also been demonstrated on non-contrast MRI. [14] In ADPKD, deployed deep learning pipelines can achieve low TKV error in clinical practice, compared to human segmentations. [15] Nevertheless, accurate segmentation of cortex, medulla, and sinus on routine non-contrast MRI remains difficult due to limited tissue contrast.

We hypothesized that training a CNN on NCE MR images paired with high-fidelity compartment labels transferred from CE MRI could learn anatomical information that compensates for limited intrinsic NCE contrast. The aim of this study was therefore to develop and validate an automated approach for renal cortex, medulla, and sinus segmentation on NCE T1-weighted MRI, and to evaluate both segmentation overlap and biomarker fidelity (volume and surface area).

## Methods

### Study Design and Participants

This retrospective, single-center study included 200 participants referred for clinically indicated abdominal MRI between January 2017 and December 2021. One examination per participant was included. Inclusion criteria required both (1) high-quality NCE T1-weighted MRI and (2) arterial-phase CE T1-weighted MRI acquired in the same session. Exclusion criteria comprised incomplete datasets, severe motion or severe non-motion artifacts (e.g. susceptibility artifacts from spine implants), genetic kidney disease, or parenchymal renal pathologies (tumors, prior partial or total nephrectomy, or more than four simple cysts per kidney). Participants were split randomly at the subject level into a development cohort (n=100; 90 training and 10 validation) and an independent test cohort (n=100) to prevent data leakage. Participants characteristics were extracted from electronic medical records. The study was conducted in accordance with the Declaration of Helsinki and approved by the local ethics committee. Written informed consent was obtained at the time of MRI acquisition.

### MRI Protocol

All participants underwent a standardized multi-sequence abdominal MRI protocol on a 1.5T scanner (MAGNETOM Avanto or MAGNETOM Aera, Siemens Healthineers, Erlangen, Germany). For model input, NCE images consisted of an axial 3D T1-weighted (T1w) chemical shift selective fat-saturated (fs) gradient-echo (GRE) acquisition acquired during 12-18 seconds breath-hold with TR 4.22 ms (range 3.50–4.22 ms) and TE 2.05 ms (range 1.41–2.09 ms). Geometric imaging parameters were field of view 400 × 300 × 200 mm (range 380–480 × 212.5– 365.6 × 180–291.2 mm), acquisition matrix 384 × 288 (range 384–416 × 204–312), and 80 slices (range 72–112). Slice thickness was 2.5 mm (range 2.5–2.8 mm) with no slice gap. The median in-plane resolution was 1.04 × 1.04 mm (range 0.91–1.25 mm), corresponding to a median voxel size of 1.04 × 1.04 × 2.5 mm.

For reference segmentation, arterial-phase CE imaging used an axial 3D T1w fs GRE sequence with the same readout and geometric parameters, acquired approximately 20–30 s after intravenous administration of a gadolinium-based contrast agent (0.2 mmol/kg gadobutrol [Gadovist®, Bayer AG, Leverkusen, Germany]; injection rate 2.0 mL/s; saline flush 25.0 mL). Arterial-phase timing was determined using a test-bolus technique (2.0 mL gadobutrol administered as the test bolus), with acquisition triggered based on the individual contrast arrival time.

### Ground Truth Segmentation and Label Transfer

Manual CE reference masks (ground truth) for renal cortex, medulla, and sinus were delineated on arterial-phase CE MRI by board-certified radiologists (2-5 years of experience in abdominal or genitourinary MRI). Segmentation was performed slice-by-slice in the axial plane, with cross-referencing in coronal and sagittal planes to ensure anatomical consistency. A second board-certified radiologist reviewed all segmentations and disagreements were resolved by consensus.

To obtain voxel-level reference on the NCE domain, the manual CE reference masks were rigidly co-registered separately for the left and right kidneys to the corresponding NCE images using SPM12 (Wellcome Centre for Human Neuroimaging, UCL; MATLAB). Rigid-body registration was performed using a normalized cross-correlation similarity metric (cost function: NCC). The optimization used a two-stage multi-resolution strategy with sampling separations (sep) of 4 mm and 2 mm (sep = [4 2]) and Gaussian smoothing of 7 mm full-width at half maximum (fwhm) applied during estimation (fwhm = [7 7]). Convergence was controlled by SPM’s standard 12-element tolerance vector, and the initial transformation parameters were set to the identity. For label transfer, CE substructure masks were temporarily merged into a whole-kidney mask, aligned to a manually generated whole-kidney reference mask on the NCE images (also created by the radiologists), and then split back into cortex, medulla, and sinus masks after transformation (Figure 1). All registrations were visually inspected. In cases of insufficient alignment, manual rigid corrections were applied before analysis.

**Figure 1.**
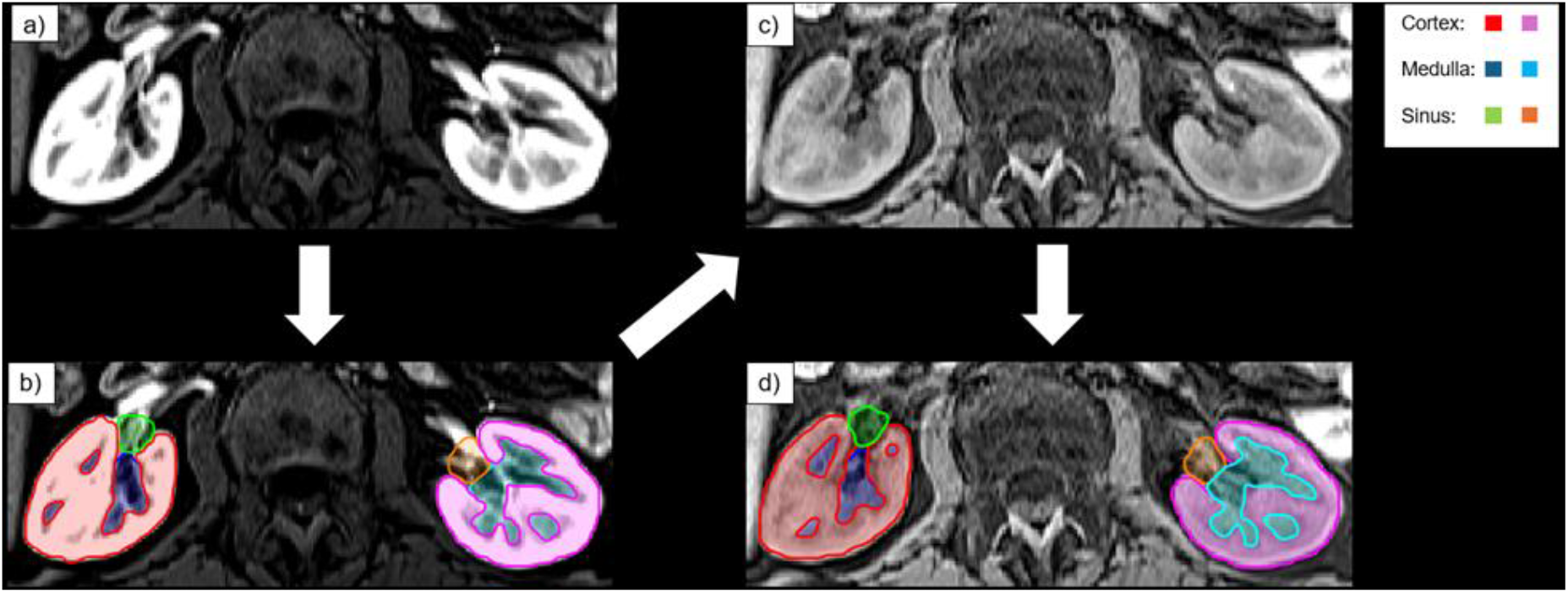
Ground Truth Segmentation and CE-to-NCE Label Transfer. (a) Axial arterial-phase contrast enhanced (CE) MRI. (b) Manually delineated masks of renal cortex, medulla, and sinus on the CE image. (c) Corresponding axial non-contrast enhanced (NCE) MRI. (d) Substructure masks after rigid CE-to-NCE co-registration.

**Figure 2.**
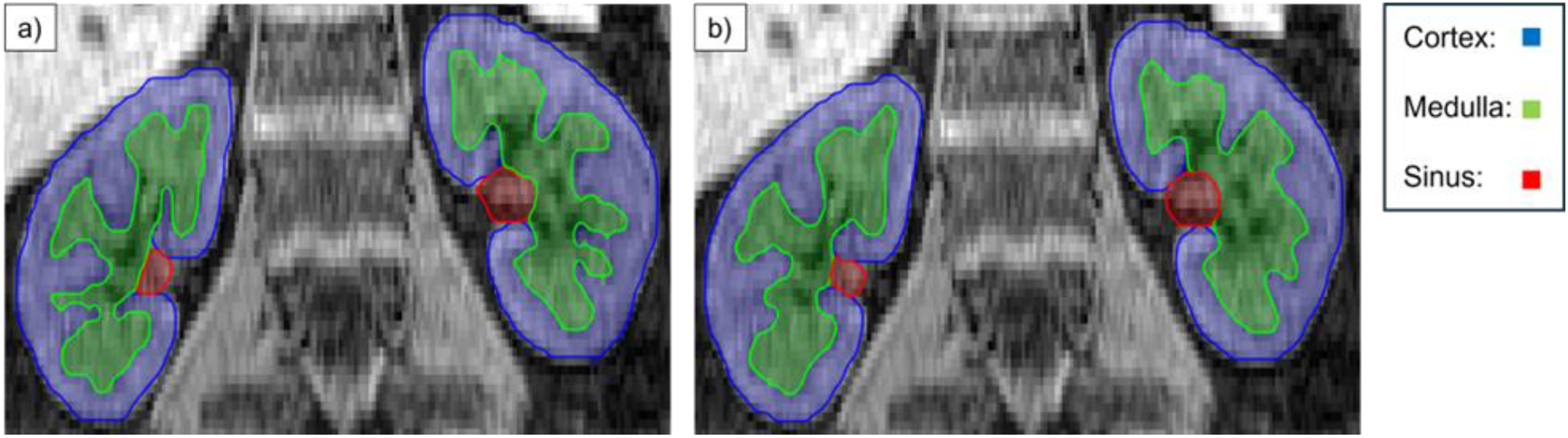
Ground truth and automated segmentation. (a) Transferred contrast enhanced masks on non-contrast enhanced image. (b) Convolutional neural network predicted masks.

### Model Development

We designed a hierarchical 3D CNN model inspired by the Deep Neural Patchworks (DNP) framework. [16] DNP is a multi-scale, patch-based segmentation approach that combines coarse contextual understanding with fine-scale local refinement, enabling accurate predictions on large 3D volumes while maintaining memory-efficient inference. Similar automated kidney segmentation pipelines have been applied in large-scale cohorts including the UK Biobank and the German National Cohort (NAKO). [13, 17]

### Biomarker Computation

To suppress potential false-positive predictions outside the kidneys, the two largest spatially connected components were retained and assigned to left and right kidneys. Within each kidney, three compartments were available: cortex, medulla, and sinus. For each compartment, volume (mL) and surface area (mm^2^) were computed from both manual CE reference masks and model predictions. Volumes were computed as voxel count multiplied by voxel volume. Surface area was computed using a voxel-face counting approach (summing exposed voxel faces at the compartment boundary and scaling by in-plane and through-plane pixel spacing). Kidney-level aggregates (left, right) were defined as the sum of cortex, medulla, and sinus. Total kidney was the sum of left and right kidneys. Bilateral compartment measures (cortex, medulla, sinus) were computed by combining both sides.

### Evaluation Metrics and Statistical Analysis

Segmentation overlap was assessed using the Dice similarity coefficient for whole kidney and each compartment, comparing the model predictions against the transferred CE reference masks on NCE images, reported as mean ± SD. For each biomarker, association between predicted and ground-truth values was quantified using Pearson correlation (r) and Spearman rank correlation (rho), with two-sided p-values. Absolute error was summarized by mean absolute error (MAE). Agreement was assessed using Lin’s concordance correlation coefficient (CCC) and Bland-Altman analysis (bias and 95% limits of agreement) (Figure 3). As an intensity-based quality control, median and IQR intensities within predicted and ground-truth compartment masks were compared on NCE images.

**Figure 3.**
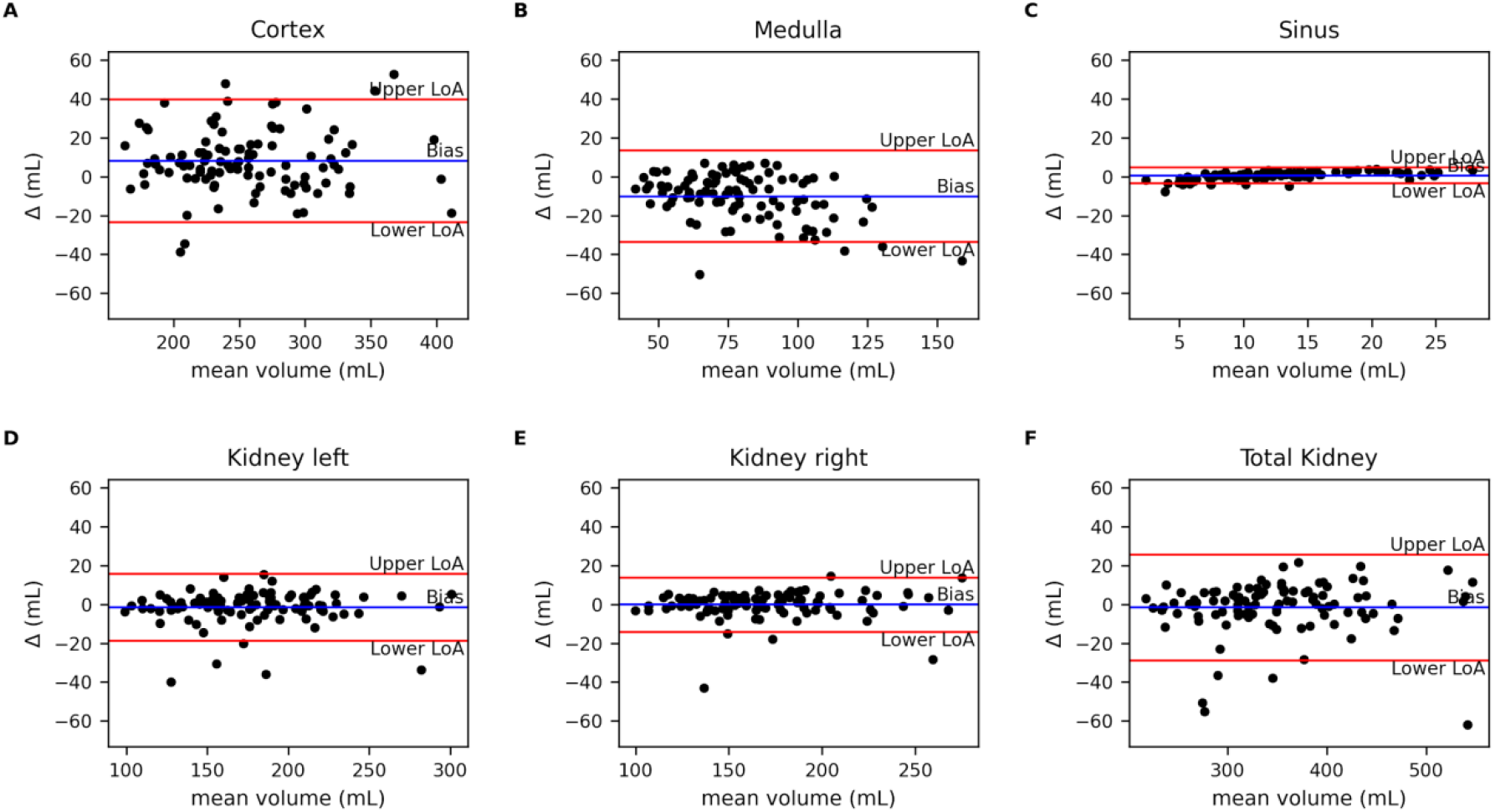
Bland–Altman Analysis of CNN-Predicted vs. Ground Truth Renal Volumes. Bland–Altman analysis of Convolutional neural network vs. ground truth volumes for bilateral cortex (A), medulla (B), sinus (C), left kidney (D), right kidney (E), and total kidney (F). Δ = prediction − ground truth (mL); positive values indicate overestimation. Blue line: bias; red lines: 95% limits of agreement (LoA)

For error analysis, we examined whether Dice performance was associated with inter-compartment intensity separation on NCE images. Relative separation **Δ**I (A, B) between compartments A and B was defined as **Δ**I = (A-B)/((A+B)/2), using median intensities within ground-truth masks. For multiple comparisons in the error analysis, p-values were adjusted using false discovery rate (FDR); adjusted q-values <0.05 were considered significant.

## Results

### Cohort Characteristics and Ground Truth Biomarkers

The mean age of the cohort was 57.3 ± 15.2 years, 44.5% of participants were female and the mean eGFR was 93.49 ± 25.37 mL (Table 1). Based on ground truth segmentations, mean total kidney volume (TKV) was 349.39 ± 75.29 mL; cortex, medulla, and sinus volumes were 251.84 ± 53.52 mL, 85.03 ± 25.21 mL, and 12.52 ± 4.86 mL, respectively (Table 3). Here the cortex accounted for approximately 72.1% of renal volume, followed by the medulla (24.3%) and sinus (3.6%). Compared with women, men had higher renal volumes (men vs. women: TKV 396.31 ± 65.63 vs. 302.36 ± 51.25 mL; cortex 281.20 ± 47.13 vs. 221.66 ± 39.69 mL; medulla 99.84 ± 25.53 vs. 70.87 ± 16.39 mL; sinus 15.26 ± 4.79 vs. 9.82 ± 3.23 mL).

**Table 1.**
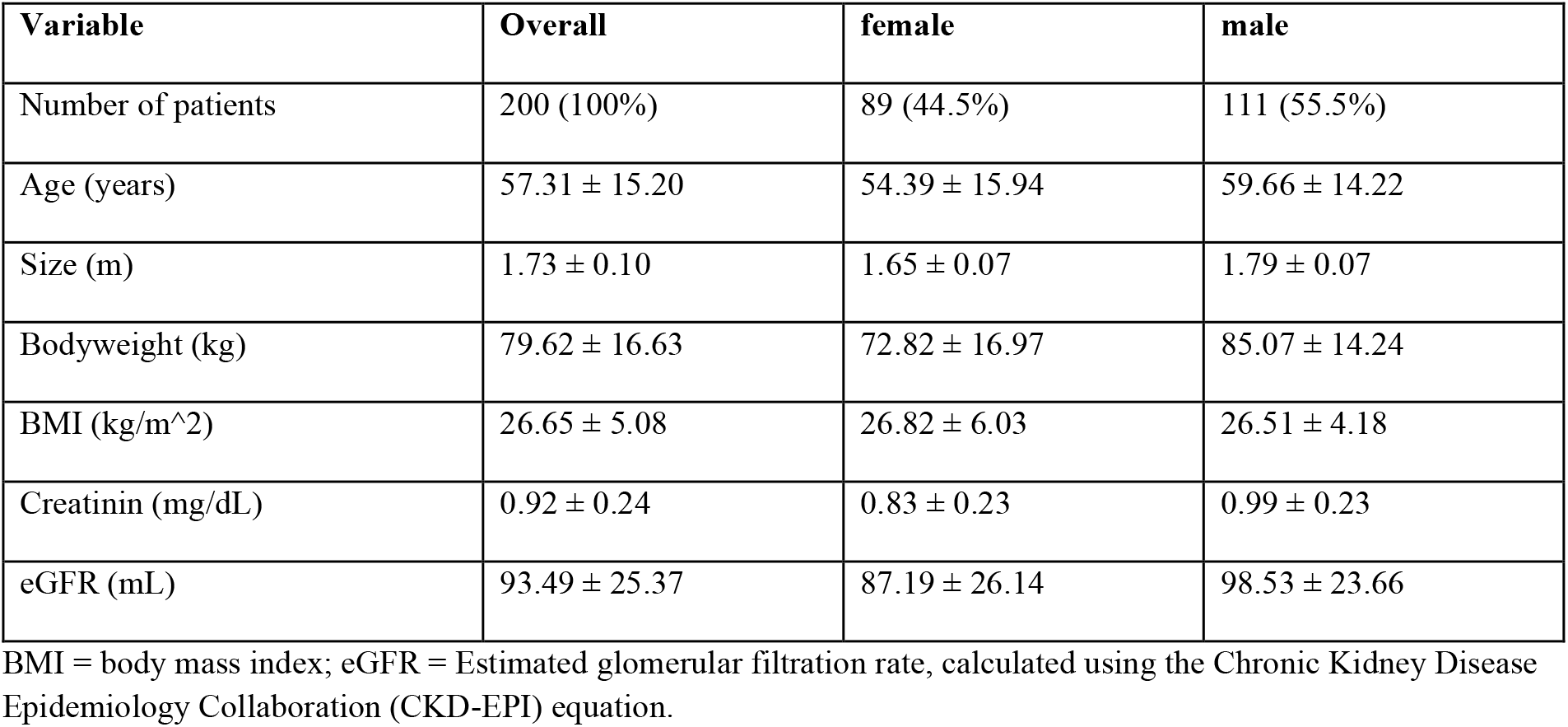
Participants characteristics of entire cohort.

### Segmentation Overlap

Whole-kidney segmentation achieved high Dice similarity on NCE MRI (left 0.950 ± 0.027; right 0.953 ± 0.027). Compartment Dice was lower but followed a consistent ranking — highest for cortex (left 0.878 ± 0.045; right 0.877 ± 0.048), intermediate for medulla (left 0.786 ± 0.077; right 0.775 ± 0.087), and lowest and most variable for sinus (left 0.790 ± 0.142; right 0.776 ± 0.134) (Table 2). Performance was broadly symmetric between left and right kidneys.

**Table 2.** Segmentation performance (Dice similarity coefficient).

| Structure | Dice (mean ± SD) |
| --- | --- |
| Cortex left | 0.878 ± 0.045 |
| Cortex right | 0.877 ± 0.048 |
| Medulla left | 0.786 ± 0.077 |
| Medulla right | 0.775 ± 0.087 |
| Sinus left | 0.790 ± 0.142 |
| Sinus right | 0.776 ± 0.134 |
| Kidney left | 0.950 ± 0.027 |
| Kidney right | 0.953 ± 0.027 |

Values are reported as mean ± standard deviation (SD)

### Biomarker Fidelity on NCE MRI

Volume biomarkers derived from predicted masks were strongly associated with ground truth across all structures (Pearson r 0.874 to 0.983; Spearman rho 0.849 to 0.985; all p < 0.001) (Table 3). Kidney-level volumes showed the highest agreement (CCC 0.976 to 0.983). At the total-kidney level, predicted TKV closely matched the ground-truth distribution (mean 347.83 vs 349.39 mL; SD 76.05 vs 75.29 mL), with minimal bias (-1.56 mL, about -0.45% of the mean) and MAE 8.76 mL (about 2.51% of mean). The 95% limits of agreement were -28.81 to 25.69 mL (about ±7.8% of mean). Left and right kidney segmentation performance was similarly well calibrated (Table 3). Rank-order agreement was also high for kidney-level volumes (Spearman rho up to 0.985), supporting reliable relative stratification.

**Table 3.** Volume biomarkers (mL): association, error, and agreement (ground truth vs prediction).

| Structure | GT mean<br>± SD (mL) | Pred<br>mean ±<br>SD (mL) | Pearson r | Spearman<br>ρ | MAE<br>(mL) | CCC | Bias (95%<br>CI) | 95% LoA |
| --- | --- | --- | --- | --- | --- | --- | --- | --- |
| Cortex left | 128.59 ±<br>29.23 | 131.68 ±<br>29.43 | 0.948 | 0.938 | 7.16 | 0.943 | 3.09 [1.21,<br>4.97] | [-15.48,<br>21.66] |
| Medulla<br>left | 43.44 ±<br>13.25 | 38.69 ±<br>10.74 | 0.884 | 0.876 | 5.9 | 0.803 | -4.76 [-<br>6.00, -<br>3.51] | [-17.03,<br>7.52] |
| Sinus left | 6.93 ± 2.71 | 7.18 ± 3.44 | 0.950 | 0.943 | 0.97 | 0.921 | 0.25 [0.01,<br>0.49] | [-2.12,<br>2.62] |
| Kidney left | 178.97 ±<br>40.86 | 177.55 ±<br>41.17 | 0.977 | 0.973 | 5.39 | 0.976 | -1.42 [-<br>3.17, 0.34] | [-18.75,<br>15.92] |
| Cortex<br>right | 123.25 ±<br>26.76 | 128.19 ±<br>27.21 | 0.943 | 0.942 | 7.59 | 0.928 | 4.94 [3.14,<br>6.74] | [-12.86,<br>22.74] |
| Medulla<br>right | 41.59 ±<br>13.32 | 36.20 ±<br>10.72 | 0.874 | 0.849 | 6.26 | 0.777 | -5.39 [-<br>6.69, -<br>4.09] | [-18.19,<br>7.41] |
| Sinus right | 5.59 ± 2.35 | 5.90 ± 2.87 | 0.919 | 0.887 | 0.93 | 0.895 | 0.31 [0.08,<br>0.54] | [-1.98,<br>2.59] |
| Kidney<br>right | 170.43 ±<br>37.89 | 170.29 ±<br>38.44 | 0.983 | 0.978 | 4.45 | 0.982 | -0.14 [-<br>1.56, 1.28] | [-14.17,<br>13.89] |
| Cortex<br>bilateral | 251.84 ±<br>53.52 | 259.87 ±<br>53.99 | 0.955 | 0.946 | 13.45 | 0.944 | 8.03 [4.82,<br>11.23] | [-23.62,<br>39.68] |
| Medulla<br>bilateral | 85.03 ±<br>25.21 | 74.89 ±<br>20.28 | 0.882 | 0.870 | 11.59 | 0.785 | -10.15 [-<br>12.53, -<br>7.76] | [-33.73,<br>13.44] |
| Sinus<br>bilateral | 12.52 ±<br>4.86 | 13.08 ±<br>6.11 | 0.953 | 0.948 | 1.71 | 0.924 | 0.56 [0.14,<br>0.97] | [-3.52,<br>4.63] |
| Total<br>kidney | 349.39 ±<br>75.29 | 347.83 ±<br>76.05 | 0.983 | 0.985 | 8.76 | 0.983 | -1.56 [-<br>4.32, 1.20] | [-28.81,<br>25.69] |

CCC = Lin's concordance correlation coefficient; CI = confidence interval; GT = ground truth; LoA = limits of agreement; MAE = mean absolute error; Pred = prediction; SD = standard deviation

At the compartment level, cortex and sinus volumes tended to be modestly overestimated, while medulla volumes were underestimated (Table 3). This resulted in a small shift in predicted compartment fractions (cortex 74.7% vs 72.1% in ground truth; medulla 21.5% vs 24.3%; sinus 3.8% vs 3.6%). Notably, sinus volume agreement remained high (bilateral CCC 0.924) despite greater dispersion in sinus Dice (Tables 2 and 3), indicating that biomarker fidelity was not fully captured by overlap metrics.

Surface area biomarkers showed only moderate correlation and substantially lower agreement compared with volume. Total kidney surface area was systematically underestimated (bias - 23787 mm^2^, about -19.9% of mean) with low concordance (CCC 0.428) and wide limits of agreement (Table 4). Nevertheless, correlations for surface area were moderate (Pearson r 0.73 to 0.80), suggesting that relative ranking may still be feasible in some settings despite poor interchangeability of absolute values.

**Table 4.** Surface area biomarkers (mm^2^): association, error, and agreement (ground truth vs prediction).

| Structure | GT mean<br>± SD<br>(mm<sup>2</sup>) | Pred<br>mean ±<br>SD (mm<sup>2</sup>) | Pearson r<br>(p) | Spearman<br>ρ (p) | MAE<br>(mm<sup>2</sup>) | CCC | Bias (95%<br>CI) | 95% LoA |
| --- | --- | --- | --- | --- | --- | --- | --- | --- |
| Cortex left | 38811.77 ±<br>10293.35 | 32852.09 ±<br>6080.89 | 0.800 | 0.813 | 6028.06 | 0.561 | -5959.68 [-<br>7258.30, -<br>4661.06] | [-<br>18787.42,<br>6868.05] |
| Medulla<br>left | 18886.96 ±<br>8030.13 | 13421.00 ±<br>3251.52 | 0.700 | 0.762 | 5511.9 | 0.348 | -5465.95 [-<br>6696.96, -<br>4234.94] | [-<br>17625.81,<br>6693.91] |
| Sinus left | 2576.53 ±<br>863.05 | 2266.75 ±<br>817.77 | 0.679 | 0.691 | 475.95 | 0.635 | -309.78 [-<br>443.65, -<br>175.91] | [-1632.15,<br>1012.58] |
| Kidney left | 60275.26 ±<br>18671.04 | 48539.84 ±<br>9686.07 | 0.754 | 0.790 | 11830.74 | 0.47 | -11735.42<br>[-<br>14321.06, -<br>9149.77] | [-<br>37276.29,<br>13805.46] |
| Cortex<br>right | 38245.93 ±<br>9865.42 | 32263.28 ±<br>5597.73 | 0.761 | 0.765 | 6073.32 | 0.511 | -5982.65 [-<br>7308.57, -<br>4656.73] | [-<br>19080.05,<br>7114.75] |
| Medulla<br>right | 18858.13 ±<br>8001.20 | 13047.61 ±<br>2962.79 | 0.632 | 0.664 | 5856.86 | 0.281 | -5810.52 [-<br>7108.80, -<br>4512.24] | [-18634.9,<br>7013.86] |
| Sinus right | 2268.04 ±<br>838.13 | 2009.52 ±<br>703.35 | 0.659 | 0.639 | 424.99 | 0.614 | -258.53 [-<br>387.22, -<br>129.83] | [-1529.72,<br>1012.67] |
| Kidney<br>right | 59372.10 ±<br>18189.45 | 47320.41 ±<br>8898.45 | 0.709 | 0.711 | 12180.79 | 0.413 | -12051.69<br>[-<br>14717.90, -<br>9385.49] | [-<br>38388.34,<br>14284.95] |
| Cortex<br>bilateral | 77057.70 ±<br>19334.70 | 65115.37 ±<br>10988.53 | 0.781 | 0.796 | 11982.68 | 0.521 | -11942.33<br>[-<br>14472.40, -<br>9412.26] | [-<br>36934.21,<br>13049.55] |
| Medulla<br>bilateral | 37745.09 ±<br>15413.31 | 26468.61 ±<br>5848.74 | 0.677 | 0.729 | 11306.75 | 0.306 | -11276.47<br>[-<br>13703.78, -<br>8849.16] | [-<br>35253.31,<br>12700.36] |
| Sinus<br>bilateral | 4844.57 ±<br>1535.28 | 4276.26 ±<br>1465.53 | 0.713 | 0.714 | 835.99 | 0.665 | -568.31 [-<br>794.25, -<br>342.37] | [-2800.11,<br>1663.49] |
| Total<br>kidney | 119647.36<br>± 35397.17 | 95860.25 ±<br>17531.98 | 0.733 | 0.759 | 23844.11 | 0.428 | -23787.11<br>[-<br>28846.72, -<br>18727.51] | [-<br>73765.66,<br>26191.44] |

CCC = Lin's concordance correlation coefficient; CI = confidence interval; GT = ground truth; LoA = limits of agreement; MAE = mean absolute error; Pred = prediction; SD = standard deviation

### Intensity-Based Quality Control and Error Analysis

Within-mask intensity distributions on NCE images were similar between predicted and ground-truth masks (Table 5). Cortical median intensity showed small positive bias (bilateral Delta median +0.68), whereas medullary and sinus medians were slightly lower in predictions (bilateral Delta median -3.80 and -2.14, respectively). IQR differences were modest across compartments.

**Table 5.** Intensity-based quality control on NCE MRI (within-mask medians and IQR).

| Structure | GT Median | Pred Median | Bias ΔMedian (95% CI) | MAE ΔMedian | GT Median IQR | Pred Median IQR | Bias ΔIQR (95% CI) | MAE ΔIQR |
| --- | --- | --- | --- | --- | --- | --- | --- | --- |
| Cortex left | 89.0<br>(80.0, 99.5) | 89.0<br>(80.8, 100.3) | 0.82<br>[0.60, 1.04] | 0.96 | 18.5<br>(16.0, 22.0) | 19.5<br>(16.8, 22.3) | 0.82<br>[0.55, 1.09] | 1.18 |
| Cortex right | 87.0<br>(75.8, 101.0) | 88.0<br>(76.0, 100.0) | 0.53<br>[0.32, 0.74] | 0.81 | 18.5<br>(16.0, 23.0) | 20.0<br>(16.0, 24.0) | 0.72<br>[0.45, 0.99] | 1.2 |
| Medulla left | 66.0<br>(59.8, 76.0) | 62.5<br>(55.8, 73.0) | -3.78 [-4.52, -3.04] | 3.78 | 19.0<br>(14.8, 26.0) | 21.0<br>(15.8, 28.0) | 1.46<br>[0.67, 2.24] | 2.02 |
| Medulla right | 66.5<br>(56.8, 74.5) | 62.0<br>(53.8, 70.3) | -3.82 [-4.37, -3.27] | 3.82 | 17.0<br>(14.0, 22.0) | 19.0<br>(15.0, 23.0) | 1.01<br>[0.45, 1.57] | 1.71 |
| Sinus left | 50.5<br>(41.5, 60.0) | 48.5<br>(39.5, 57.0) | -2.84 [-4.45, -1.23] | 3.22 | 28.0<br>(22.0, 33.3) | 31.5<br>(23.8, 37.0) | 2.55<br>[1.74, 3.35] | 3.69 |
| Sinus right | 51.0<br>(41.8, 60.0) | 49.0<br>(40.0, 58.0) | -1.44 [-2.64, -0.23] | 2.8 | 27.0<br>(21.0, 32.3) | 30.0<br>(24.9, 36.0) | 3.52<br>[2.59, 4.46] | 3.99 |
| Cortex bilateral | 87.2<br>(79.4, 99.3) | 88.3<br>(79.8, 100.3) | 0.68<br>[0.50, 0.85] | 0.82 | 18.5<br>(16.0, 21.6) | 19.8<br>(16.5, 22.5) | 0.77<br>[0.55, 0.99] | 1.1 |
| Medulla bilateral | 66.3<br>(58.9, 75.13) | 63.8<br>(54.4, 70.1) | -3.80 [-4.40, -3.20] | 3.8 | 18.5<br>(14.5, 24.0) | 20.0<br>(15.5, 24.6) | 1.23<br>[0.60, 1.86] | 1.71 |
| Sinus bilateral | 50.3<br>(42.9, 60.5) | 48.3<br>(40.4, 56.6) | -2.14 [-3.11, -1.17] | 2.72 | 27.8<br>(21.4, 32.1) | 31.3<br>(23.9, 35.5) | 3.04<br>[2.36, 3.71] | 3.57 |

Comparison of NCE signal intensities between ground truth (GT) and model predictions (Pred). For each compartment, median and the median of interquartile range (IQR) are reported for GT and Pred, along with the corresponding bias (Pred – GT) and mean absolute error (MAE).

Segmentation Dice correlated with relative inter-compartment intensity separation (ΔI) computed from ground-truth compartment intensities (Table 6). For example, medulla Dice correlated strongly with cortex-medulla separability (Pearson r 0.631, q < 0.001), and sinus Dice correlated with both cortex-sinus and medulla-sinus separability (Pearson r 0.523 and 0.437, q < 0.001).

These associations remained similar when analyzed separately for each side.

**Table 6.**
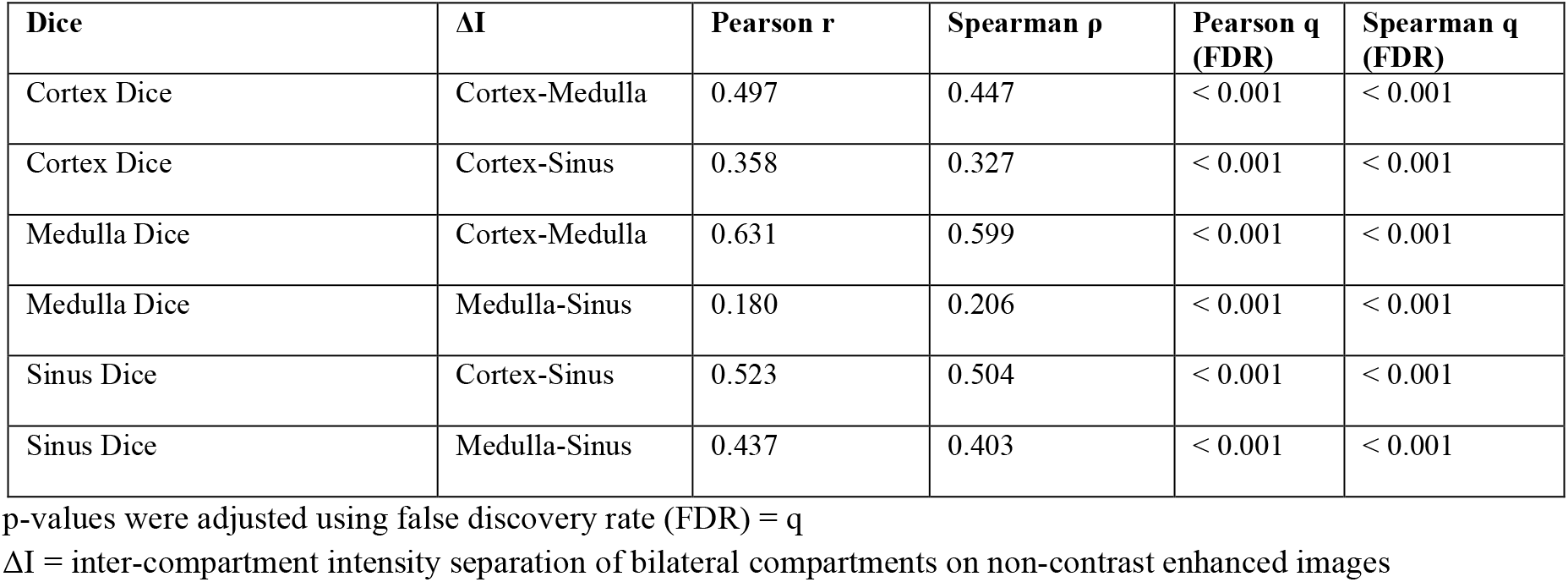
Correlation between Dice and relative inter-compartment intensity separation (ΔI), pooled across sides.

## Discussion

This study demonstrates that CE-supervised label transfer can enable accurate, fully automated extraction of renal imaging biomarkers from routine non-contrast T1-weighted MRI. Whole-kidney segmentation achieved Dice ∼0.95 for both sides, and kidney-level volumetry was not only strongly correlated with ground truth but also well calibrated, with minimal bias and preserved population distribution. These properties are critical for longitudinal monitoring and for cohort-scale analyses where absolute calibration and distributional fidelity matter as much as overlap.

Our whole-kidney Dice scores are comparable in magnitude to prior population-scale kidney segmentation work; for example, automated segmentation of parenchymal kidney volume in UK Biobank neck-to-knee body MRI reported Dice around 0.95. [13] Likewise, our TKV error is in the range reported by deployed ADPKD segmentation pipelines used for automated volumetry. [15]

A key finding is that biomarker fidelity was not fully captured by voxel-level overlap metrics for smaller compartments. Sinus Dice exhibited the greatest dispersion, yet sinus volume agreement remained high (bilateral CCC 0.924). This illustrates a common pitfall of relying solely on overlap: small or topologically complex compartments can show large Dice variability from small boundary shifts, while still providing clinically relevant volumetry. Consequently, biomarker-first evaluation (correlation, error, agreement) is necessary when the downstream goal is quantitative phenotyping rather than pixel-perfect boundary replication.

At the compartment level, the model showed a systematic pattern: cortical volumes were modestly overestimated and medullary volumes were underestimated, leading to a small shift in predicted volume fractions (cortex up, medulla down). This aligns with the error analysis, where medulla Dice correlated strongly with cortex-medulla intensity separability on NCE images. The result is physiologically plausible: corticomedullary contrast is reduced on routine NCE T1-weighted acquisitions compared with arterial-phase CE imaging, and the model appears to revert to an anatomical prior when separability is low. In practice, this implies that cortex-medulla phenotypes derived from NCE-only pipelines should be interpreted with awareness of potential systematic bias, and that calibration or sequence optimization may further improve compartment balance.

Surface area biomarkers showed substantially poorer agreement than volume, with systematic underestimation and wide limits of agreement. Surface area is inherently more boundary sensitive than volume, and our voxel-face counting method discretizes the boundary. Both factors amplify the impact of small segmentation differences. While moderate correlations suggest that relative ordering may still be possible for certain applications, absolute surface areas from NCE-derived masks should not be considered interchangeable with ground truth in their current form. Future work could evaluate mesh-based surface estimation with consistent resampling and boundary regularization to reduce discretization effects.

The ability to derive kidney and compartment volumes from contrast-free MRI is attractive for longitudinal follow-up and large cohort studies. Kidney size metrics and volumetry (including TKV in ADPKD) are recognized structural biomarkers in kidney imaging research and clinical studies. [3-6, 18] In addition, cortical volume is a key imaging component in approaches combining imaging with biopsy-derived glomerular density to estimate nephron number. [7] Because gadolinium-based contrast agents are commonly avoided or used with caution in advanced CKD due to safety and deposition concerns, non-contrast alternatives can facilitate broader implementation. [9-10] Moreover use of contrast medium is not feasible in large population-based studies e.g. NAKO.

This study has limitations. First, it is a retrospective single-center dataset with a specific acquisition protocol; external validation across scanners, field strengths, and institutions is needed. Second, ground truth in the NCE domain relied on rigid registration of CE labels, which can introduce local mismatch, particularly near the renal hilum or with motion between acquisitions. Third, surface area estimation used a voxel-face counting approach that may not generalize across resolutions. Fourth, the cohort predominantly comprised participants with preserved renal function and without major structural distortion. While this reduces anatomical confounding and supports the internal validity of the CE-to-NCE transfer framework, performance in advanced CKD or markedly remodeled kidneys remains to be established and warrants external validation. Finally, the evaluation treated left and right kidneys as separate units for several analyses. While appropriate for side-specific performance reporting, this may introduce within-subject correlation that is not modeled in simple pooled statistics.

In conclusion, CE-supervised knowledge transfer using a hierarchical patch-based CNN enables accurate and well-calibrated kidney volumetry from routine non-contrast T1-weighted MRI. The approach supports contrast-free extraction of clinically relevant renal imaging biomarkers, while highlighting that compartment surface areas and cortex-medulla balance remain sensitive to intrinsic contrast and boundary uncertainty.

## Conclusion

CE-supervised knowledge transfer using a hierarchical patch-based CNN enabled accurate and well-calibrated kidney volumetry from routine non-contrast T1-weighted MRI (total kidney volume MAE ∼2.5% with minimal bias). While cortex, medulla, and sinus overlap metrics were lower than whole kidney, volume biomarkers remained strongly associated with ground truth.

## Data Availability

All data produced in the present study are available upon reasonable request to the authors.

## Abbreviations

ADPKD: autosomal dominant polycystic kidney disease
BA: Bland-Altman
CCC: concordance correlation coefficient
CE: contrast-enhanced
CKD: chronic kidney disease
CNN: convolutional neural network
CRISP: Consortium for Radiologic Imaging Studies of Polycystic Kidney Disease
DNP: Deep Neural Patchworks
FDR: false discovery rate
fwhm: full-width at half maximum
IQR: interquartile range
LoA: limits of agreement
MAE: mean absolute error
MRI: magnetic resonance imaging
NCE: non-contrast-enhanced
sep: separations
TKV: total kidney volume
T1w: T1-weighted.

## Data Availability

Available from the corresponding author on reasonable request.

